# Does a postpartum “Green Star” family planning decision aid for adolescent mothers reduce decisional conflict? : A quasi-experimental study

**DOI:** 10.1101/2022.03.10.22272184

**Authors:** Stella E. Mushy, Eri Shishido, Shigeko Horiuchi

## Abstract

**Aim:** To our knowledge, there are still no studies in Tanzania regarding decision aids on long-acting reversible contraception. We evaluated the effects of our *postpartum “Green Star” family planning decision aid* on decisional conflict, knowledge, satisfaction, and uptake of long-acting reversible contraception among pregnant adolescents in Tanzania.

**Methods:** We used a facility-based quasi-experimental design with control. The participants were purposively recruited and randomly assigned (intervention, n = 33; control, n = 33). The intervention received the routine family planning counseling and decision aid. The control received only the routine family planning counseling. The primary outcome was change in decisional conflict measured using the validated Decisional Conflict Scale (DCS). The secondary outcomes were knowledge, satisfaction, and contraception uptake. We hypothesized that pregnant adolescents who use the decision aid will have a lower DCS score.

**Results:** We recruited 66 pregnant adolescents; 62 completed the study. Participants in the intervention had a lower mean difference score in the DCS than participants in the control (intervention: -24.7 [SD 7.99] vs. control: -11.6 [SD 10.9], t = -5.53, p < 0.001). The mean difference score in knowledge was significantly higher in the intervention than in the control (intervention: 4.53 [SD 2.54] vs. control: 2.0 [SD 1.45], t = 4.88, p < 0.001). The mean score of satisfaction was significantly higher in the intervention than in the control (intervention: 100 [SD 0.0] vs. control: 55.8 [SD 30.7], t = 8.112, p < 0.001). Choice of contraception was significantly higher in the intervention [29 (45.3%)] than in the control [13 (20.3%)] (x^2^ = 17.73, p < 0.001).

**Conclusion:** The *postpartum “Green Star” family planning decision aid* was useful as it lowered decisional conflict, improved knowledge and satisfaction with decision making, and enhanced contraception uptake. The decision aid demonstrated positive applicability and affordability for pregnant adolescents in Tanzania.

## Introduction

Adolescent pregnancies remain a global public health concern, with the highest rate occurring in developing countries. Across the world, about 16 million girls aged 15-19 years are estimated to give birth each year. [1] Most of these pregnancies are unintended, [1, 2] that is, unplanned or unwanted pregnancies. [3]

According to the latest Tanzania Demographic Health Survey, the adolescent population (10-19 years old) comprises 23% of the total population (44.9 million). Most adolescents are sexually active before the age of 15 years, and the number of childbearing adolescents aged 15-19 years has gradually increased from 23% in 2010 to 27% in 2016. Moreover, the adolescent fertility rate has increased from 116 to 132 for every 1000 girls between 2010 and 2016, and almost 56.7% of adolescents have begun childbearing at the age of 19 years.[4]

Adolescent pregnancies remain a major national problem as well as a critical health and social priority in Tanzania.[5, 6] One-quarter (24%) of the pregnancies among young women are unintended.[7] The country statistics show that 21% of young women’s pregnancies occur among adolescents aged 15-19 years, of which 18.1% are first births and 2.9% are second or later births.[4]

Most women who start childbearing in their teenage years have more children and shorter birth spacing. Furthermore, most of their births are unintended compared with women who began childbearing when they were 20 years or older.[8] Subsequent pregnancies during the teen years compound the economic, physical, and psychological consequences of adolescent mothers.[8, 9] Failure to complete high school and impaired economic sufficiency are significantly associated with repeated pregnancy during adolescent age.[10]

Various factors increase the risk of unintended adolescent pregnancies and rapid repeat pregnancies (RRPs). These include inconsistent use of contraception, lack of knowledge about immediate use of family planning after birth, lack of communication between teens and their healthcare providers, fear of side effects, and myths and misconceptions about family planning.[11-13] The main barriers hindering a female youth from going to family planning services are weight gain, reduced sexual desire, irregular bleeding, infertility, and abnormal vaginal discharge.[11]

To prevent early pregnancies, countries across the world, including Tanzania, have implemented sexual and reproductive health education, enhanced negotiation skills building, and improved awareness and accessibility to modern contraceptives among adolescents.[14] However, most interventions focus on preventing first pregnancies with fewer efforts on preventing subsequent unintended pregnancies during the teen years. Thus, most adolescents continue to experience RRP despite the availability and accessibility of family planning methods. The majority (78%) of non-first pregnancies among adolescent mothers in Tanzania experience RRP within 3 months after birth, with over half of the births (51%) occurring in the second year from the last birth.[13] This is very alarming and calls for researchers, healthcare providers, and policymakers to exert considerable and sustained effort into supporting pregnant adolescents in the planning and timing of their subsequent pregnancies.

Delaying the second birth will provide a better chance for an adolescent mother to mature psychologically and biologically. Ultimately, this increases her opportunities to complete high school, make plans for the future, and develop other training skills.[15] Delaying the second birth will also reduce the risks of premature births, stillbirths, and newborn morbidity and mortality.[16]

One of the primary and essential strategies for reducing high-risk pregnancies, which often occur too early and frequently, is the effective use of family planning.[17] The use of long-acting reversible contraception immediately after childbirth (i.e., intrauterine copper devices and implants) is considered a high-impact intervention that reduces the risks associated with adolescent pregnancy.[18] These methods do not rely on daily, weekly, or monthly use, and have higher continuation and satisfaction rates than short-acting reversible contraception.[18] Research findings from a previous study indicated that adolescent mothers who initiated long-acting reversible contraception after delivery had a lower risk of RRP than adolescent mothers who started short-acting reversible contraception or who did not use any family planning methods.[19]

Despite significant gains in the training of healthcare providers, distribution of family planning commodities, and provision of quality of care, the postpartum contraceptive uptake among adolescent mothers in Tanzania remains critically underutilized. Only 12.2% of teenage mothers use postpartum contraceptives within three months after delivery, with the largest proportion using injectables followed by pills.[4] Both intrauterine copper devices and implants are highly effective in preventing pregnancy, and they all last for several years and are easy to use.[1, 18] For these reasons, pregnant adolescents need comprehensive information about all the contraception options available in their country to help them decide which contraceptive they can use. In the case of pregnant adolescents in Tanzania, this involves clarifying their values and beliefs and what is important to them, particularly regarding intrauterine copper devices and implants being the only locally available long-acting reversible contraception methods.

On the other hand, decision aids in clinical settings have been beneficial as they inform and educate patients about the available treatment options, which help reduce decisional conflicts.[20] Recently, decision aids have been widely used to provide health information to patients and the public.[21] As intrauterine copper devices and implants have almost equal effectiveness and are long-lasting devices, the choice of pregnant adolescents will depend on their preferences and lifestyle. Pregnant adolescents must clarify their values by appraising the benefits and harms of each option and weighing attributes that are personally important to them when making a choice.

Decision aids on family planning counseling have been shown to have a positive outcome. [22, 23] However, to our knowledge, there are still no studies on decision aids that focus on long-acting reversible contraception methods to improve family planning uptake by adolescent mothers immediately after childbirth. To avoid the long-term physiological, psychological, and economic consequences adolescent mothers must face from unintended RRP owing to non-contraception use, we developed a decision aid named *postpartum “Green Star” family planning decision aid*. We borrowed the name “Green Star” from the Kiswahili word “Nyota ya Kijani”, a well-known word to most people in Tanzania. The Tanzanian government launched the National Family Planning project in 1992 and used the Kiswahili word “Nyota ya Kijani” as the project name. The project focused on sensitizing women of reproductive age to use artificial family planning methods to properly space their pregnancy and limit the number of their children. The *postpartum “Green Star” family planning decision aid* that we newly developed includes updated evidence-based information on intrauterine copper devices and implants to guide pregnant adolescents decide regarding the best option for using long-acting reversible contraception methods based on their preference. Its relevance lies in the guidance provided to pregnant adolescents in using artificial family planning methods to properly space and reduce their pregnancy, similarly to the principle of “Nyota ya Kijani.”

The *postpartum “Green Star” family planning decision aid* was assessed for its feasibility and found to be practical, useful, and acceptable in addressing the underutilization of postpartum family planning methods.[11] In the current study, we evaluated the effects of the *postpartum “Green Star” family planning decision aid* on Decisional conflict, Knowledge, Satisfaction, and Uptake of contraception among pregnant adolescents in Tanzania. We hypothesized that pregnant adolescents who are using the *postpartum “Green Star” family planning decision aid* will have a lower decision conflict scale (DCS) score than pregnant adolescents who are not using the decision aid (control group), and that knowledge and satisfaction scores will be higher in pregnant adolescents using the decision aid than in pregnant adolescents who are not using the decision aid (control group).

## Materials and methods

### Study design, setting, and participants

This study used a quasi-experimental design with a control. The participants were pregnant adolescents attending antenatal care services at Mkuranga and Kisarawe district hospitals in the Pwani region. The inclusion criteria were pregnant adolescents who were between 15-19 years, in their 28 gestation weeks, planning to deliver at the hospital where they are attending antenatal care services, and willing and provided consent to participate. The exclusion criteria were pregnant adolescents who were receiving family planning education from other programs.

Adolescents were selected as the study population because they have a high risk of RRP and they critically underutilize different types of modern contraception compared with women above 20 years. [13] The occurrence of RRP in adolescents more severely worsens the health risks and compounds the socioeconomic inequality than that in women above 20 years because involvements in education and work training are repeatedly delayed and are less likely to be attained.

The Pwani region is among the regions in Tanzania with the highest childbearing rates. Thirty percent (30%) of pregnant women in the Pwani region are usually women aged 15-19 years. [4] Mkuranga and Kisarawe are among the 6 districts of the Pwani region in Tanzania. The selected district hospitals are government-run public health facilities that also serve low-income populations. Thus, the turn-up of participants in these district hospitals was expected to be higher than that in other district hospitals and private health facilities. The study sites (i.e., antenatal clinic, labor, and postpartum wards) were co-located within the facilities. Mkuranga district hospital was assigned as the interventional hospital whereas Kisarawe district hospital was assigned as the control hospital.

The sample size in the current study design was based on the minimum sample size suggested by the Central Limit Theorem of 30 participants in each group.[24] We considered a dropout rate of 10%, making the number of participants in each group 33. Thus, the total number of participants recruited in this study was 66.

The Ethics Boards of St. Luke’s International University (20-A091), Muhimbili University of Health and Allied Sciences (MUHAS-REC-1-2020-076), and National Institute of Medical Research approved this study. This study was registered in the Clinical Trials Registry of University Hospital Information Network in Japan (UMIN000043085). We also obtained permission from the regional and district administrative officers, district medical officers, and medical staff in charge of the selected hospitals for data collection. The study researchers explained the purpose, scope, and importance of the study to each participant. The participants who voluntarily agreed to participate in the study provided both oral and written informed consents.

Purposive sampling was used to select the study participants. Enrolment of the study participants in both groups run concurrently, and the study sites were in the different districts which reduced data contamination risk. Eligible participants in both groups received three education sessions before giving birth. Three education sessions offered at Time 1, 2, and 3 were considered sufficient for the participants to understand the methods and address all the barriers preventing them from using family planning. For each education session, each participant in the intervention group first received the routine family planning counseling offered. Then, this was followed by individual face-to-face family planning counseling education using the contents of the *postpartum “Green Star” family planning decision aid*. It took about 30-40 minutes for the research assistant (RA) to present all the contents to each participant. Every participant received the 10-page *postpartum “Green Star” family planning decision aid* and brought it home for further reading and as reference when needed. The participants in the control group only received a routine family planning counseling offered at each antenatal clinic visit. Each participant received three education sessions as in the intervention group. The differences in the education session contents between the intervention group and the control group are shown in **Box 1**. The study participants were followed up three times (at 32 gestation week [Time 2], at 36/38 gestation weeks [Time 3], and at the discharge date after delivery [Time 4]) before completing the study. Recruitment, enrolment and data for the intervention and control groups were carried out from 1^st^ March 2021 to the end on September 2021.

#### Box 1.

**Differences in education session contents between intervention and control groups**

**Box 1. Differences in education session contents between intervention and control groups**
| Education session contents | Intervention group<sup>1,2</sup> | Control group<sup>2</sup> |
| --- | --- | --- |
| Description of the family planning methods | ✓ | ✓ |
| Making an explicit decision | ✓ |  |
| Information on long-acting reversible contraception options | ✓ | ✓ |
| Information on other family planning methods | ✓ | ✓ |
| Side effects and disadvantages | ✓ | ✓ |
| Benefits and advantages | ✓ | ✓ |
| Comparative tables of long-acting reversible contraception options | ✓ |  |
| Values clarification exercise | ✓ |  |

1: *postpartum “Green Star” family planning decision aid* 2: *Routine family planning guide*

Two RAs (AM and CM) who were certified midwives recruited the participants and conducted the study in close coordination with the lead researcher (SM). The criteria for selecting the RAs were experience in family planning counseling, ability to conduct intervention studies, and working in an antenatal clinic. Each RA received two days of training separately to avoid data contamination. The training focused on how to clearly explain the study purpose; who to recruit; and what, when, and how they should educate the participants. The RAs were also trained on when and how they should conduct a close follow-up of the participants to minimize lost to follow-up. The RAs were taught on the types of activities to be carried out at each visit, as well as the kinds of information to be collected at each visit. The RAs were also trained on the appropriate time to obtain oral and written informed consents from the study participants.

### Development of the postpartum “Green Star” family planning decision aid

We developed the *postpartum “Green Star” family planning decision aid* by initially identifying the research gap, target population, and study objectives to address the research problem on adolescent pregnancy. We carefully focused and examined previously published studies when determining the study objectives.[21, 23, 25-27] Currently, there are limited publications describing the use of decision aids in reducing decision-making conflicts regarding the utilization of long-acting reversible contraception methods.

Thereafter, we identified the individual needs of the participating pregnant adolescents by reviewing a previous study that looked into barriers to the utilization of family planning among female youths in Dar es Salaam, Tanzania.[11] The individual needs of the participants included inadequate knowledge, particularly of long-acting reversible contraception methods. We found that female teens have several misbeliefs about contraception methods and how to participate in decision-making. Most female teens could not decide on their own without involving their sexual partners.

The content, design, and arrangement of the developed prototype decision aid were based on the Ottawa Patient Decision Aid Development eTraining,[28] International Patient Decision Aid Standards Collaboration checklist,[29] Theory of Planned Behavior,[30] Health Belief Model,[31] Social Cognitive Theory,[32] current clinical guides for family planning counseling for providers,[1, 33] and findings from previous studies on the benefits and side effects of the different options, satisfaction and continuation rates, and fertility return.[34-40] The prototype decision aid has four components based on the Ottawa Patient Decision Aid Development eTraining guide: i) know how to make a decision with conviction; ii) understand the characteristics of the decision; iii) clarify what is important to you, and iv) make the decision. We shared the prototype decision aid with three experts, namely, our research supervisor and two midwives, all with extensive years of experience in maternal and child health and in developing decision aids. The aim of sharing the prototype decision aid was to receive comments on the comprehensibility and usefulness of the prototype decision aid, which we incorporated in the modified and improved *postpartum “Green Star” family planning decision aid*.

We then previously carried out a feasibility study of the prototype decision aid to assess its practicality, usefulness, and acceptability as perceived by pregnant adolescents and healthcare providers.[41] In our previous feasibility study, we interviewed 6 healthcare providers and 12 pregnant adolescents. We asked the study participants questions about the length, flow, and comprehensibility of the prototype decision aid, and if this tool can meet the decision-making needs of pregnant adolescents. Based on the feedback from the study participants, we revised the prototype decision aid. Several comments were given such as the need to rearrange the flow of the contents, and to change some Kiswahili and English words to reduce ambiguity. The prototype decision aid was designed for home use as a preconsultation support for the option of using long-acting reversible contraception. However, both the healthcare providers and the pregnant adolescents suggested that this tool should be implemented by healthcare providers. After incorporating all the comments and suggestions provided, we developed the second version of the decision aid. This second version of the decision aid was shared among developer experts for final check and approval. We received and integrated several comments and suggestions from the developer experts. Finally, we developed the third and final version of the decision aid and named it *postpartum “Green Star” family planning decision aid*. This decision aid was then assessed for its effects on Decisional conflict, Knowledge, Satisfaction, and Uptake of long-acting reversible contraception.

## Measurements

### Primary outcome: Decisional conflict

Decisional conflict was assessed at Time 1 (28 gestational weeks) and at Time 4 (within 2 days after childbirth) using the decisional conflict scale (DCS). The DCS is a 10-item self-report questionnaire that measures a patient’s uncertainty about what treatment option to choose and the factors associated with the uncertainty (e.g., lack of information, myths and misconception, and lack of support).[42] The DCS has 4 subscales: Informed, Clarity, Support, and Uncertainty). Items were answered using a 3-point Likert scale (0 = “Yes”; 2 = “unsure”; 4 = “no”) with scores ranging from 0 to 100. A higher score meant a higher decisional conflict and uncertainty, and vice versa.[42] As we adapted items in the DCS, we conducted Cronbach’s Alpha test to check for internal consistency of the DCS items, and we obtained 0.848 which was higher than the commonly recommended value of 0.6. The results indicated that a set of items in the DCS were reliable (i.e., closely related as a group).

### Secondary outcomes

The secondary outcomes were Knowledge, Satisfaction with decision-making, and Uptake of long-acting reversible contraception. Knowledge was assessed at Time 1 and Time 3 (36-38 gestational weeks). We created a knowledge questionnaire to test the participants’ knowledge of long-acting reversible contraception. We adapted questions from different reports,[11, 33-35, 38-39, 43] which included advantages, disadvantages, side effects, myths, and misconceptions commonly existing in the community. The knowledge questionnaire had 7 questions each with a score of 1 for a total of 7 scores. The questions required a “yes” or “no” response. A score of 7 indicated that a participant is knowledgeable about long-acting reversible contraception and vice versa. Satisfaction was only assessed at Time 4. We assessed satisfaction using an effective decision-making subscale of the DCS that had 4 items with 3 responses (4 = “yes”, 2 = “unsure”, 0 = “no”) with scores ranging from 0 to 100. A higher score indicated a higher satisfaction with decision-making and vice versa. The decision on which option to use following childbirth was only assessed at Time 4 using 1 question that asked, “which option do you prefer?”

### Demographic data

The information collected as part of the questionnaire included Age, Parity, Highest education level, Occupation, and Marital status.

### Data processing and analysis

Data were descriptively analyzed using IBM SPSS Statistics version 24.0. Data cleaning was performed using frequency distribution tables to confirm if the data were correctly entered. To ensure the validity and reliability of the data results, each variable was manually cross-checked and corrections were made when necessary to avoid altering the statistical analysis. The Shapiro-Wilk test was conducted, and the histogram distributions were checked. Descriptive analysis was used to analyze the demographic information of the participants to determine frequencies and percentages of their distribution within groups. The chi-square test was conducted to analyze data and to observe the distribution within the group for the ordinal and between groups for categorical data. The analysis was based on calculating the mean score differences of the selected variables at Time 1, Time 2, Time 3, and Time 4 between groups. The independent sample t-test was performed to compare the mean scores of DCS, Knowledge, and Satisfaction between groups. The statistical tests were performed with a two-sided 5% level of significance.

## Results

### Flow of the study

Data were collected for 7 months from early March to the end of September 2021. The participant flow diagram is shown in **Fig 1**. A total of 80 pregnant adolescents were eligible, but only 66 gave their written informed consent and were included in the study with 33 participants in each group. Two participants, one from the intervention and one from the control group were lost to follow-up and were excluded from the analysis. Therefore, a total of 64 participants were included for analysis, each group with a total of 32 participants.

**Fig 1.**
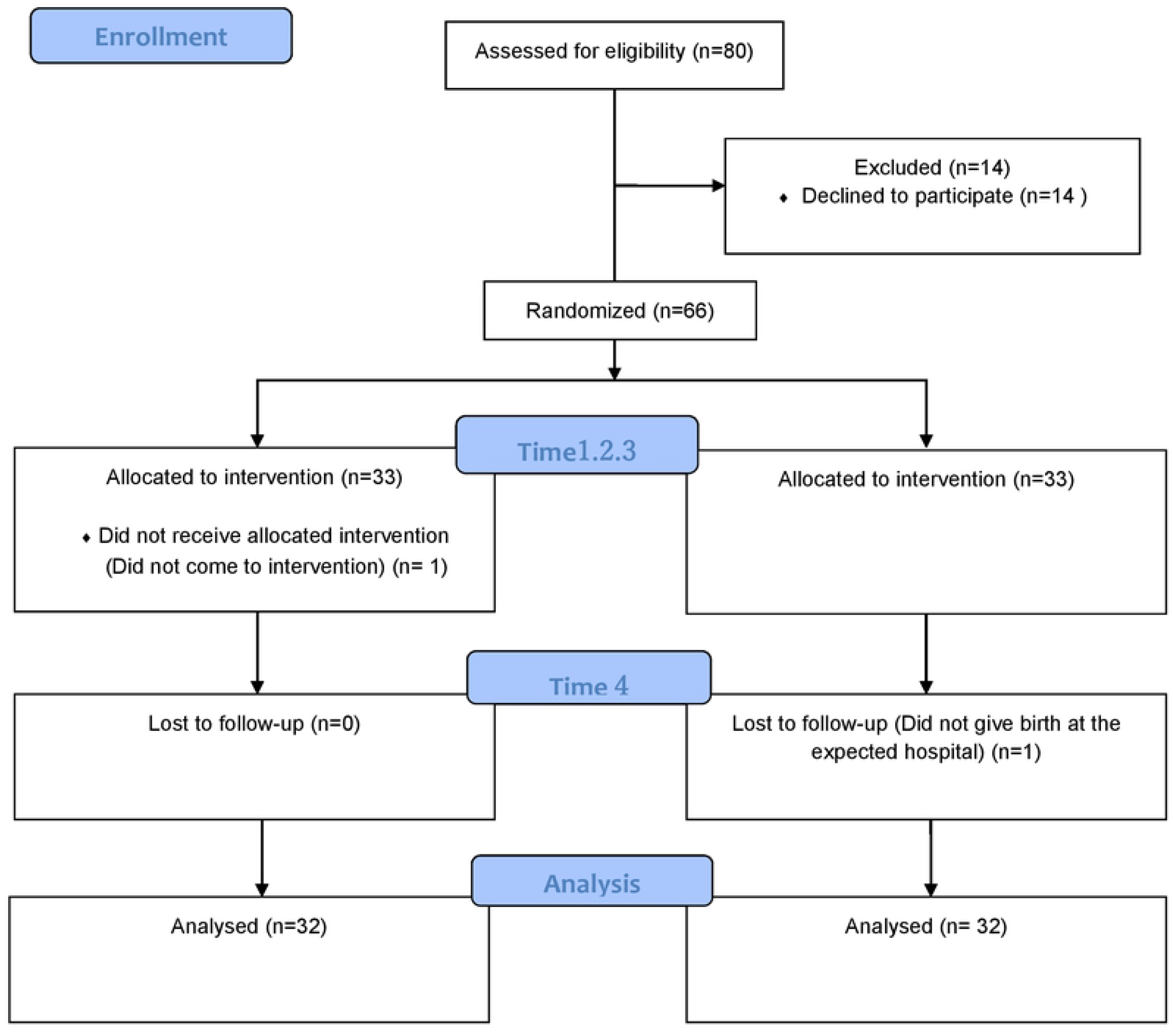
Flow diagram of study participants.

### Sociodemographic characteristics of the study participants

Details of the sociodemographic information of the study participants are shown in **Table 1**. The mean age of the study participants in the intervention group was 17.5 (1.29) years, whereas that in the control group was 18.0 years (SD 0.71) [age range, 15-19 years]. Marital status and occupation showed a significant difference between the 2 groups (p < 0.001). The ratio of single mothers was higher in the intervention group [23 (71.9%)] than in the control group [7 (21.9%)]. Similarly, the ratio of self-employed mothers was higher in the intervention group [27 (84.4%)] than in the control group [8 (25%)].

**Table 1.** Sociodemographic characteristics of the study participants.

| | Intervention group<br>(n = 32) | Control group<br>(n = 32) | t | $\chi^2$ | p-value |
| --- | --- | --- | --- | --- | --- |
| **Age (years)** |  |  |  |  |  |
| mean | 17.5 (SD 1.29) | 18.0 (SD 0.71) | -2.15 |  | 0.030 |
| **Marital status (%)** |  |  |  |  |  |
| Single | 23 (71.9) | 7 (21.9) |  | 19 | < 0.001 |
| Married | 9 (28.1) | 17 (53.1) |  |  |  |
| Cohabiting | 0 | 8 (25.0) |  |  |  |
| **Gravidity (%)** |  |  |  |  |  |
| 1 | 31 (96.9) | 29 (90.6) |  | 1.07 | 0.302 |
| 2 | 1 (3.1) | 3 (9.4) |  |  |  |
| **Highest education level (%)** |  |  |  |  |  |
| Primary | 27 (84.4) | 27 (84.4) |  | 0.48 | 0.788 |
| Secondary | 3 (9.4) | 4 (12.5) |  |  |  |
| None | 2 (6.2) | 1 (3.1) |  |  |  |
| **Occupation (%)** |  |  |  |  |  |
| Employee | 0 | 1 (3.1) |  |  | < 0.001 |
| Self-employed | 27 (84.4) | 8 (25.0) | 45.98 |  |  |
| Housewife | 1 (3.1) | 20 (62.5) |  |  |  |
| Do not work | 4 (12.5) | 3 (9.4) |  |  |  |

### Decisional conflict

The DCS mean scores at Time 1 and Time 4 in the intervention and control groups are shown in **Table 2**. The total DCS mean score at Time 1 was significantly lower in the intervention group than in the control group (intervention group: 65.00 [SD 19.1] vs. control group: 77.80 [SD 18.4], t = -2.72, p = 0.008). The observed mean scores in the DCS subscales “Support” (p = 0.723) and “Uncertainty” (p = 0.548) were not significantly different between the 2 groups at Time 1. However, at Time 4, there was a significant difference in the total DCS mean score between the intervention group and the control group (intervention group: 3.13 [SD 4.7] vs. control group: 48.5 [SD 29.6], t = -8.55, p < 0.001). Additionally, the mean scores of the all 4 subscales (i.e., Informed, Clarity, Support, and Uncertainty) in the DCS between the 2 groups at Time 4 showed a significant difference (p < 0.001). The mean difference score in the DCS (Time 4 minus Time 1) was significantly lower in the intervention group than in the control group (intervention group: -24.7 [SD 7.99] vs. control group: -11.6 [SD 10.9], t = -5.53, p < 0.001). The mean difference scores of all 4 subscales (i.e., Informed, Clarity, Support, and Uncertainty) in the DCS were significantly lower in the intervention group than in the control group. These results support the hypothesis that the decision-making conflict score will be lower in the intervention group than in the control group.

**Table 2.**
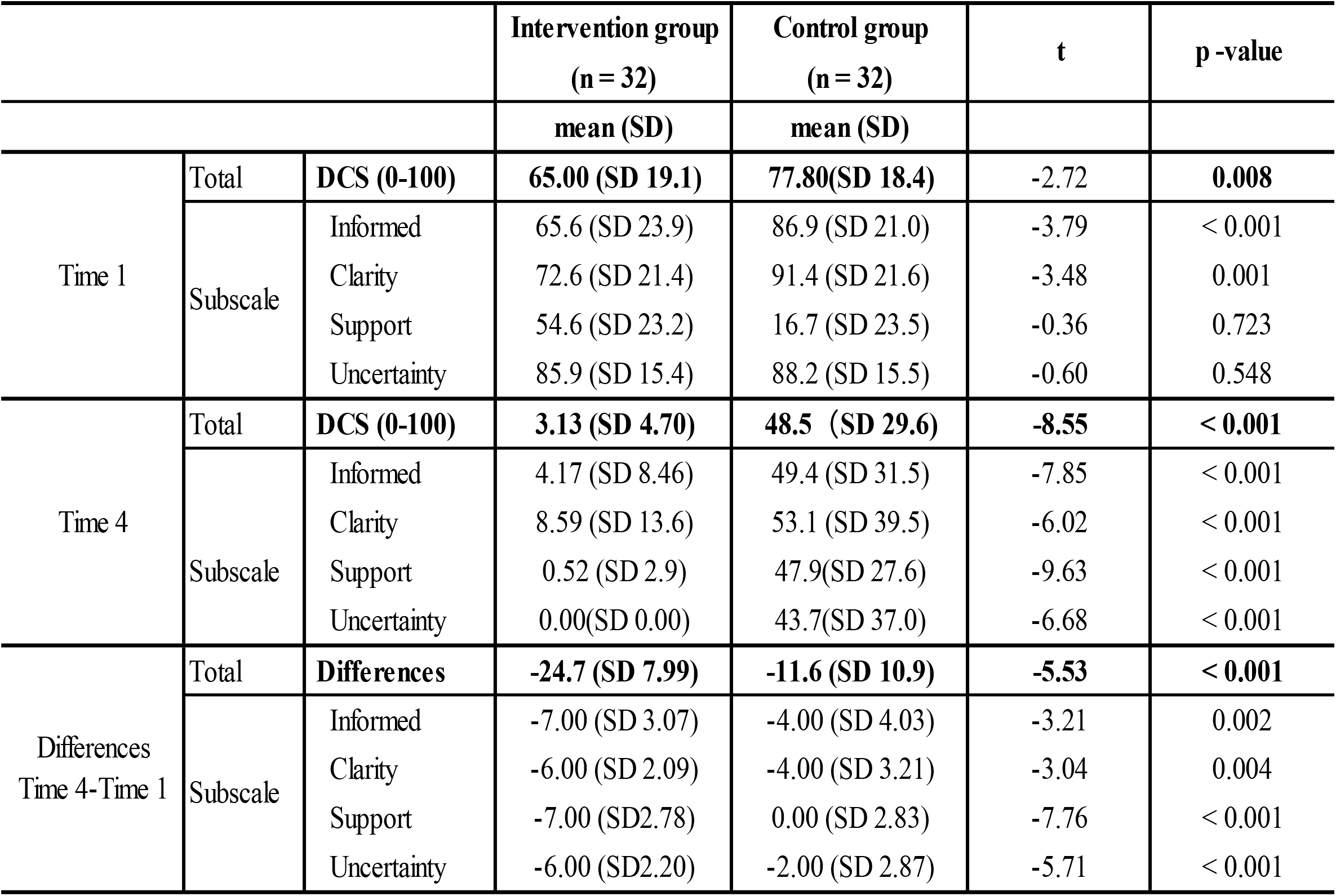
Mean Scores of the Decisional Conflict Scale.

|  |  |  | Intervention group<br>(n = 32) | Control group<br>(n = 32) | t | p -value |
| --- | --- | --- | --- | --- | --- | --- |
|  |  |  | mean (SD) | mean (SD) |  |  |
| Time 1 | Total | **DCS (0-100)** | **65.00 (SD 19.1)** | **77.80(SD 18.4)** | -2.72 | **0.008** |
|  | Subscale | Informed | 65.6 (SD 23.9) | 86.9 (SD 21.0) | -3.79 | < 0.001 |
|  |  | Clarity | 72.6 (SD 21.4) | 91.4 (SD 21.6) | -3.48 | 0.001 |
|  |  | Support | 54.6 (SD 23.2) | 16.7 (SD 23.5) | -0.36 | 0.723 |
|  |  | Uncertainty | 85.9 (SD 15.4) | 88.2 (SD 15.5) | -0.60 | 0.548 |
| Time 4 | Total | **DCS (0-100)** | **3.13 (SD 4.70)** | **48.5 (SD 29.6)** | **-8.55** | **< 0.001** |
|  | Subscale | Informed | 4.17 (SD 8.46) | 49.4 (SD 31.5) | -7.85 | < 0.001 |
|  |  | Clarity | 8.59 (SD 13.6) | 53.1 (SD 39.5) | -6.02 | < 0.001 |
|  |  | Support | 0.52 (SD 2.9) | 47.9(SD 27.6) | -9.63 | < 0.001 |
|  |  | Uncertainty | 0.00(SD 0.00) | 43.7(SD 37.0) | -6.68 | < 0.001 |
| Differences<br>Time 4-Time 1 | Total | **Differences** | **-24.7 (SD 7.99)** | **-11.6 (SD 10.9)** | **-5.53** | **< 0.001** |
|  | Subscale | Informed | -7.00 (SD 3.07) | -4.00 (SD 4.03) | -3.21 | 0.002 |
|  |  | Clarity | -6.00 (SD 2.09) | -4.00 (SD 3.21) | -3.04 | 0.004 |
|  |  | Support | -7.00 (SD2.78) | 0.00 (SD 2.83) | -7.76 | < 0.001 |
|  |  | Uncertainty | -6.00 (SD2.20) | -2.00 (SD 2.87) | -5.71 | < 0.001 |

### Multiple linear regression for predicting DCS score

Multiple linear regression was performed to predict the DCS score based on Age, Occupation, and Marital status at Time 1, Time 4, and at the time Differences (Time 4 minus Time 1). Occupation was treated as a dummy variable. Age was the only variable that showed a significant relationship in the control group, both at Time 1 (*β* = 0.455, p = 0.015) and at Time 4 (*β* = 0.506, p = 0.006). However, Age did not show a significant relationship at the time Differences (Time 4 minus Time 1). As for the intervention group, none of the variables showed a significant relationship with the DCS scores (**Table 3**).

**Table 3.** Multiple linear regression for predicting DCS score.

|  | Study arms | Items | Beta | p-value |
| --- | --- | --- | --- | --- |
| Time 1 | Intervention group | Age | 0.292 | 0.120 |
|  |  | Occupation (dummy) | -0.115 | 0.532 |
|  | Control group | Age | 0.455 | **0.015** |
|  |  | Occupation (dummy) | -0.308 | 0.090 |
| Time 4 | Intervention group | Age | -0.227 | 0.217 |
|  |  | Occupation (dummy) | 0.305 | 0.099 |
|  | Control group | Age | 0.506 | **0.006** |
|  |  | Occupation (dummy) | -0.092 | 0.597 |
| Time 4-Time 1 | Intervention group | Age | -0.333 | 0.073 |
|  |  | Occupation (dummy) | 0.182 | 0.317 |
|  | Control group | Age | 0.240 | 0.211 |
|  |  | Occupation (dummy) | 0.108 | 0.572 |

### Contraceptive knowledge, satisfaction and uptake

At Time 1, the results showed no significant difference in the Knowledge mean score between the 2 groups (intervention group: 1.84 [SD 1.98] vs. control group: 2.34 [SD 1.61], t = -1.1, p = 0.274). At Time 3, the Knowledge mean score was significantly higher in the intervention group than in the control group (intervention group: 6.38 [1.60] vs. control group: 4.34 [1.82], t = 4.733, p < 0.001). The mean difference score in Knowledge (Time 3 minus Time 1) in the intervention group was larger than that in the control group (intervention group: 4.53 [2.54] vs. control group: 2.00 [1.45], t = 4.88, p < 0.001). These findings support the hypothesis that the level of knowledge of long-acting reversible contraception will be higher in the intervention group than in the control group.

The mean score of Satisfaction was significantly higher in the intervention group than in the control group (intervention group: 100.00 [SD 0.0] vs. control group: 55.8 [SD 30.7], t = 8.112, p < 0.001). The proportion of “yes” responses of each item about Satisfaction was greater in the intervention group (100%) than in the control group (< 40%). The chi-square test result of each item between the 2 groups was found to be significant in all items (p < 0.001). These results support the hypothesis that the state levels of Satisfaction with decision making will be higher in the intervention group than in the control group.

The proportion of participants who decided to use long-acting reversible contraception showed significant differences between the 2 groups. The proportions of participants who “did not decide to use any option” were 3 (4.7%) in the intervention group and 19 (29.7) in the control group. However, the proportions of participants who “decided to use implant” were 29 (45.3%) in the intervention group and 13 (20.3%) in the control group (x^2^ = 17.73, p < 0.001). These results showed that all long-acting reversible contraception users only opted for implants and nobody chose an intrauterine copper device. These findings support the hypothesis that the proportion of long-acting reversible contraception uptake will be higher in the intervention group than in the control group.

### Logistic regression analysis for long-acting reversible contraception uptake

Age, Marital status, and Occupation showed no significant relationship with long-acting reversible contraception uptake in the intervention group. However, Age alone showed a significant relationship with long-acting reversible contraception uptake in the control group with a negative *β* value (**Table 4**).

**Table 4.** Logistic regression analysis for long-acting reversible contraception uptake.

| Study arms | Item | Beta | p-value | Odds | 95% CI |  |
| --- | --- | --- | --- | --- | --- | --- |
|  |  |  |  |  | Lower | Upper |
| Intervention group | Age | -0.56 | 0.45 | 0.57 | 0.13 | 2.50 |
|  | Marital status | -19.52 | 1 | 0 | 0 |  |
|  | Occupation | -20.22 | 1 | 0 | 0 |  |
| Control group | Age | -1.55 | **0.03** | 0.21 | 0.05 | 0.84 |
|  | Marital status | -0.62 | 0.32 | 0.53 | 0.16 | 1.82 |
|  | Occupation | 0.08 | 0.87 | 1.09 | 0.38 | 3.17 |

## Discussion

We evaluated the effects of our recently developed *postpartum “Green Star” family planning decision aid* on pregnant adolescents in terms of their usage of long-acting reversible contraception following childbirth. Our hypotheses were as follows: pregnant adolescents who use the *postpartum “Green Star” family planning decision aid* will have a lower DCS score than pregnant adolescents who do not use the decision aid; Knowledge and Satisfaction scores, and the proportion of contraception uptake will be higher in pregnant adolescents who receive the *postpartum “Green Star” family planning decision aid* than in pregnant adolescents who do not receive the decision aid. All of these hypotheses were supported by our results.

### Decisional conflict for adolescent mothers

Decision aids are crucial tools for use in clinical settings as they provide patients a chance to be involved in making medical decisions by weighing the risks and benefits in a balanced manner. [20] The present findings indicate that the total mean score of DCS was significantly lower in the intervention group than in the control group. The mean difference scores in the subscales of DCS were significantly lower in the intervention group than in the control group.

The present results concur with the results of a systematic review that involved 105 studies on decision aids from different areas, namely, cancer screening, prenatal complication diagnosis, immunization, and diabetic treatments. [20] These studies found that the use of decision aids more markedly reduced the mean difference in the DCS score in the intervention group (with decision aids) than in the control group (without decision aids) (MD -9.28, 95% CI: -12.2 to -6.36). The effect of decision aids on medication choice for diabetes mellitus was also assessed in 3 randomized clinical trials.[27, 44, 45] The findings of these trials showed a significantly lower mean difference in the DCS score in the intervention group than in the control group. Two clinical trials involving pregnant women have also been conducted.[25, 46] The first clinical trial assessed the effect of decision aids on the choice of pregnant women whether to have epidural anesthesia or not during labor.[25] The second randomized clinical trial evaluated the effect of decision aids on women with breech presentation at term.[46] The findings of these previous trials were similar to the findings of the present study in that the mean difference in the DCS score was significantly lower in the intervention group which received a decision aid than in the control group which received only a standardized routine care.

On the other hand, the present findings are inconsistent with the findings from a previous study that evaluated the effect of a decision aid on decision-making for the treatment of pelvic organ prolapse. [47] The previous study found that the DCS score of patients who received a decision aid and standard counseling was not significantly lower than the DCS score of patients who received only standard counseling (p = 0.566). The probable reasons include the already available pelvic organ prolapse decision aid in the setting and the regular review of information by the patients together with the clinician at the initial encounter. The observed mean difference in the DCS score in the study of Brazel et al. was very low in both the intervention and control group, indicating no significant difference between the 2 groups.[47] The results suggested that the existing method of “standard counseling” alone was sufficient in their specific type of study population.

In the present study, a significant mean difference was observed in the intervention group because there are no family planning decision aids in antenatal clinics to help patients decide on the family planning option that they would take. The unavailability of family planning decision aids occurs not only in antenatal clinics but also in other clinics such that there are no decision aids to supplement routine counseling given to patients to aid in their decision-making. A study on the experiences of diabetic patients and healthcare providers on shared decision-making conducted in Tanzania found that neither the patients nor the healthcare providers had been using decision-making aids; the patients reported that only health education tools are being used for educating them. [48]

The DCS in the present study had 4 subscales that were compared between the 2 groups. The mean difference scores for the subscales Uncertainty, Clarity, Support, and Informed were significantly lower in the intervention group than in the control group (p ≤ 0.001). These findings are consistent with the findings of previous studies.[20,45,46] The participants in the intervention group expressed their feeling of being more involved in the decision-making process as well as feeling that they received sufficient information to be able to decide compared with the participants in the control group. Nevertheless, the present findings are inconsistent with the findings of Brazel et al. [47] who found no significant mean difference scores in the subscales of the DCS between the intervention and control groups: Uncertainty (p = 0.519), Clarity (p = 0.590), Support (p = 0.413), and Informed (p = 0.718).

The multiple regression analysis of the DCS score based on Age, Marital status, and Occupation showed Age as the only variable that had a significant relationship with the DCS score in the control group at Time 1 and Time 4, but not in the intervention group. The findings further show that as Age increases, the decision conflict score also increases and vice versa. These findings indicate that if younger adolescents receive the correct information about long-acting reversible contraception at the right time, this will improve their chances of utilizing family planning methods. Regarding the lower DCS score in the intervention group, the present findings suggest that the decision aid played an important role in imparting knowledge and correcting the held myths and misconceptions of the younger adolescents.

### Knowledge, satisfaction, and uptake of long-acting reversible contraception for adolescent mothers

The mean differences in the scores of Knowledge, Satisfaction, and Uptake of long-acting reversible contraception were significantly higher in the intervention group than in the control group. In a systematic review [20] of 52 studies, 4 randomized control trials [44, 45, 46, 49] and 1 survey [50] found a significant increase in the scores of Knowledge, Satisfaction with decision making, and Choice for the treatment options in the intervention group which used a decision aid compared with the control group which used a standardized routine care. Although the present study showed an increase in contraception uptake, none of the long-acting reversible contraception users chose an intrauterine copper device as all the participants chose only implants. The main reasons for not using an intrauterine copper device might be related to individual perception factors such as myths and misconceptions, and discomfort from postpartum pain. [11]

The logistic regression analysis showed that Marital status and Occupation showed no significant relationship with knowledge and Satisfaction in the 2 groups. Only Age showed a significant relationship with long-acting reversible contraception uptake in the control group. The study participants in the control group were 21% (odds = 0.21) less likely to use long-acting reversible contraception than the study participants in the intervention group. Younger adolescents in the control group were more likely to utilize long-acting reversible contraception than older adolescents. These findings indicate that if family planning programs exert greater efforts in reaching out to pregnant adolescents and ensuring that they receive the correct information at the right time regarding postpartum family planning, its uptake will be markedly improved following childbirth.

The present findings also suggest that the decision aid played a significant role in increasing Knowledge which ultimately improved the uptake of the available and preferred long-acting reversible contraception following childbirth. The making of a choice implies that pregnant adolescents were able to learn from the *postpartum “Green Star” family planning decision aid*, and this changed their minds by correcting their held myths or misconceptions about long-acting reversible contraception. The present findings also show that receiving the correct information at the right time regarding long-acting reversible contraception improves satisfaction with decision-making.

In the present study, none of the long-acting reversible contraception users used an intrauterine copper device. This non-usage was related to the fear of expulsion and risk of infection. If an intrauterine copper device is inserted immediately after childbirth and infection prevention control measures are adhered to, then the risk of expulsion and infection would be minimal. However, the *postpartum “Green Star” family planning decision aid* did not have any information regarding the timely insertion of an intrauterine copper device. In future studies, this information will be included in the decision aid to improve intrauterine copper device uptake.

## Strengths and limitations

To our knowledge, this is the first study in Tanzania that used a postpartum family planning decision aid to assist in decision-making of pregnant adolescents regarding which long-acting reversible contraception they should use following childbirth. This means the open practicality of using the decision aid for healthcare even though the subjects are adolescents. To avoid data contamination between groups, the hospital in the intervention group was located in a different district from the hospital in the control group, that is, 2-3 hours of driving using a private transport. On the other hand, the present findings cannot be generalized, unless a randomized controlled study is conducted.

## Conclusions

To our knowledge, this is the first quasi-experimental study with a control that evaluated the effects of our recently developed *postpartum “Green Star” family planning decision aid* on pregnant adolescents’ choice of using long-acting reversible contraception. The decision aid significantly lowered the decision-making conflict, improved knowledge and satisfaction with decision-making, and enhanced the uptake of available long-acting reversible contraception. The overall findings indicate the usefulness of the *postpartum “Green Star” family planning decision aid* as it supplemented and supported patient-provider communications during family planning counseling in antenatal clinics.

## Data Availability

All relevant data are within the manuscript and its Supporting Information files.

## Acknowledgments

We would like to express our gratitude to Dr. Edward Barroga (https://orcid.org/0000-0002-8920-2607), Medical and Nursing Science Editor and Professor of Academic Writing at St. Luke’s International University for reviewing and editing the manuscript.

## Funding

This work was supported by Japan Society for the Promotion of Science (JSPS) Core-to-Core Program **(2021-2024)** (PI: Shigeko Horiuchi). The funding was used in the data collection, analysis, and in writing the manuscript.

## Competing interests

The authors declare that they do not have any competing interests associated with this study.

## Supporting information

A “Green Star” family planning decision aid

## Author Contributions

**Conceptualization:** Stella E. Mushy, Eri Shishido, Shigeko Horiuchi

**Data curation:** Shigeko Horiuchi

**Formal analysis:** Stella E. Mushy, Eri Shishido, Shigeko Horiuchi

**Funding acquisition:** Shigeko Horiuchi

**Investigation:** Stella E. Mushy, Shigeko Horiuchi

**Methodology:** Stella E. Mushy, Eri Shishido, Shigeko Horiuchi

**Project administration:** Stella E. Mushy consideration

**Resources:** Shigeko Horiuchi

**Supervision:** Stella E. Mushy, Eri Shishido, Shigeko Horiuchi

**Validation:** Eri Shishido, Shigeko Horiuchi

**Visualization:** Eri Shishido

**Writing – original draft:** Stella E. Mushy

**Writing – review & editing:** Stella E. Mushy, Eri Shishido, Shigeko Horiuchi

## Notes

### Competing Interest Statement

The authors have declared no competing interest.

### Clinical Trial

ID:UMIN000043085

### Clinical Protocols

https://center6.umin.ac.jp/cgi-open-bin/icdr_e/ctr_view.cgi?recptno=R000049189

### Author Declarations

The Ethics Boards of St. Luke?s International University (20-A091), Muhimbili University of Health and Allied Sciences(MUHAS-REC-1-2020-076), and National Institute of Medical Research approved this study. This study was registered in the Clinical Trials Registry of University Hospital Information Network in Japan (UMIN000043085).

## References

1. World Health Organization. Adolescent pregnancy: key facts (2018). [Cited 2019 Jan 5]. Available from: https://www.who.int/news-room/fact-sheets/detail/adolescent-pregnancyy.

2. Neal S, Matthews Z, Frost M, Foisted H, Camacho AV, Laski L. Childbearing in adolescents aged 12–15 in low resource countries: a neglected issue. New estimates from demographic and household surveys in 42 countries. Acta Obstet Gynecol Scand. 2012; Available from: https://doi.org/10.1111/j.1600-0412.2012.01467.x.

3. Unintended Pregnancy in the United States: Fact sheet. [Cited 2021 May 11]. Available from: https://www.guttmacher.org/sites/default/files/factsheet/fb-unintended-pregnancy-us.pdf.

4. Tanzania Demographic and Health Survey and Malaria Indicator Survey. Dar es Salaam, Tanzania. MoHCDGEC Ministry of Health CD, Gender, Elderly and Children - MoHCDGEC/Tanzania Mainland, MOH Ministry of Health - MoH/Zanzibar, NBS National Bureau of Statistics - NBS/Tanzania. OCGS Office of Chief Government Statistician - OCGS/Zanzibar, ICF. 2015 – 2016. [Cited 2019 Jun 2]. Available from: https://dhsprogram.com/pubs/pdf/FR321/FR321.pdf.

5. The United Republic of Tanzania. The National Road Map Strategic Plan II to Improve Reproductive, Maternal, Newborn, Child and Adolescent Health in Tanzania. Ministry Of Health, Community Development, Gender, Elderly, and Children. 2016 – 2020. [Cited 2019 May 7]. Available from: https://www.globalfinancingfacility.org/sites/gff_new/files/Tanzania_One_Plan_II.pdf.

6. The United Republic of Tanzania. The National Family Planning Costed Implementation Program. Ministry of Health and Social Welfare. 2010-2015. [cited 2019 Apr 3]. Available from: http://ec2-54-210-230-186.compute-1.amazonaws.com/wp-content/uploads/2014/10/NFPCIP_Amendment_NEW2.pdf.

7. Population Reference Bureau. Tanzania Youth Reproductive Health: Satisfying an unmet need for family planning. 2015. [cited 2019 July 5]. Available from: https://assets.prb.org/pdf15/unmetneed-factsheet-tanzania.pdf.

8. United Nations Population Fund. Adolescent Pregnancy: A Review of the Evidence. 2013. [cited 2019 May 1]. Available from: https://www.unfpa.org/sites/default/files/pub-pdf/ADOLESCENT%20PREGNANCY_UNFPA.pdf.

9. Briggs G, Brownell M, Ross N. Teen mothers, and socioeconomic status. The chicken-egg debate. Journal of the Association for Research on Mothering”. 2007; 9(1):62–74. [Cited 2019 June 20]. Available from: https://jarm.journals.yorku.ca/index.php/jarm/article/view/5136/4332.

10. Jones ME, Mondy LW. Lessons for prevention and intervention in adolescent pregnancy: A five-year comparison of outcomes of low programs for school-aged pregnant adolescents. J Pediatr Health Care. 1994; 8(4):152–9. Available from: https://doi.org/10.1016/0891-5245(94)90027-2

11. Mushy SE, Tarimo EAM, Fredrick MA, Horiuchi S. Barriers to the uptake of modern family planning methods among female youth of Temeke District in Dar es Salaam, Tanzania: A qualitative study. Sex Reprod Healthc. 2020; 24. Available from: https://doi.org/10.1016/j.srhc.2020.100499.

12. Darroch JE, Woog V, Bankole A, Ashford LS. Adding It Up: Costs and Benefits of Meeting the Contraceptive Needs of Adolescents, New York: Guttmacher Institute. 2016. [Cited 2020 Jan 11]. Available from: https://www.guttmacher.org/report/adding-it-meeting-contraceptive-needs-of-adolescent.

13. Cleland J, Conde-Agudelo A, Peterson H, Ross J, Tsui A. Family planning needs during the first two years postpartum in Tanzania. Contraception and health. The Lancet. 2012; 380(9837):149–56. [Cited 2020 Feb 15]. Available from: https://www.mchip.net/sites/default/files/Tanzania-PPFP.pdf.

14. Oringanje C, Meremikwu MM, Eko H, Esu E, Meremikwu A, Ehiri JE. Interventions for preventing unintended pregnancies among adolescents. Cochrane Database of Syst Rev. 2016; 2:CD005215. Available from: https://doi.org/10.1002/14651858.CD005215.pub3..

15. Alford S, Rutledge A, Huberman B. Science and success. The programs that worked to prevent subsequent pregnancy among adolescent mothers. Advocates for youth. 2009. [Cited 2020 Nov 1]. Available from: https://www.advocatesforyouth.org/wpcontent/uploads/storage//advfy/documents/sspregnancies.pdf.

16. De Jonge HCC, Azad K, Seward N, Kuddus A, Shaha S, Costello A, et al. Determinants and consequences of short birth interval in rural Bangladesh: a cross-sectional study. BMC Pregnancy Childbirth. 2014; 14:427 Available from: https://doi.org/10.1186/s12884-014-0427-6.

17. Primary health care: Report of the international conference on primary health care. Alma-Ata, USSR, 6 – 12 September 1978. Jointly sponsored by the World Health Organization and the United Nations Children’s Fund. [Cited 2019 May 7]. Available from: https://www.who.int/publications/i/item/9241800011.

18. The American College of Obstetricians and Gynecologists. Long-acting reversible contraception: implants and intrauterine devices. Obstet Gynecol. 2017; 130:251–69. [Cited 2021 May 1]. Available from: https://www.acog.org/clinical/clinical-guidance/practice-bulletin/articles/2017/11/long-acting-reversible-contraception-implants-and-intrauterine-devices.

19. Baldwin MK, Edelman AB. The effect of long-acting reversible contraception on rapid repeat pregnancy in adolescents: a review. J Adolesc Health. 2013; 52(4):47 – 53. Available from: https://doi.org/10.1016/j.jadohealth.2012.10.278.

20. Stacey D, Légaré F, Lewis K, Barry MJ, Bennett CL, Eden KB, et al. Decision aids for people facing health treatment or screening decisions. Cochrane Database of Syst Rev-Intervention. 2017. Available from: https://doi.org/10.1002/14651858.CD001431.pub5.

21. Bennett KF, Wagner VC, Robb KA. Supplementing factual information with patient narratives in the cancer screening context: a qualitative study of acceptability and preferences. 2015; 18:2032 – 2041. Available from: https://doi.org/10.1111/hex.12357.

22. Wu JP, Damnshroder LJ, Fetters MD, Zikmund-Fisher BJ, Hudson SV, Fucinari JBS, et al. A Web-based decision tool to improve contraceptive counseling for women with chronic medical conditions: Protocol for a mixed-methods implementation study. 2018; 7(4). Available from: https://doi.org/10.2196/resprot.9249.

23. Kim YM, Davila C, Tellez C, Kols A. Evaluation of the World Health Organization’s family planning decision-making tool: improving health communication in Nicaragua. Patient Education and Counseling. 2007; 66(2):235 – 242. Available from: https://doi.org/10.1016/j.pec.2006.12.007.

24. Ganti A (2019). Central Limit Theorem. Oxford Executive MBA. [Cited 2021 August 13]. Available from: https://www.investopedia.com/terms/c/central_limit_theorem.asp#:∼:text=Key%20Takeaways,The%20central%20limit%20theorem%20(CLT)%20states%20that%20the%20distribution%20of,for%20the%20CLT%20to%20hold.

25. Shishido E, Osaka W, Henna A, Motomura Y, Horiuchi S. Effect of a decision aid on the choice of pregnant women whether to have epidural anesthesia or not during labor. PLoS One. 2020; 15(11). Available from: https://doi.org/10.1371/journal.pone.0242351.

26. Osaka W, Nakayama K. Effect of a decision aid with patient narratives in reducing decisional conflict in choice for surgery among early-stage breast cancer patients: A three-arm randomized controlled trial. Patient Educ Couns. 2017; 100(3):550 – 562. Available from: https://doi.org/10.1016/j.pec.2016.09.011.

27. Brown S, Lumley J. Satisfaction with care in labor and birth: A survey of 790 Australian women. Birth Issues in Prenatal Care. 1994; 21(1):4 – 13. Available from: https://doi.org/10.1111/j.1523-536x.1994.tb00909.x.

28. Ottawa Patient Decision Aid Development eTraining (ODAT). Patient Decision Aids. [Cited 2021 01 2]. Available from: https://decisionaid.ohri.ca/index.html.

29. The International Patient Decision Aid Standards (IPDAS) Collaboration. IPDAS Criteria for Judging the Quality of Patients Decision Aids. 2005. [Cited 2021 01 2]. Available from: http://ipdas.ohri.ca/IPDAS_checklist.pdf.

30. Ajzen I. The theory of planned behavior. Organizational Behavior and Human Decision Processes. 1991; 50(2):179 – 211. Available from: https://doi.org/10.1016/0749-5978(91)90020-T.

31. Green EC, Murphy E. Health belief model. In: Wiley Blackwell Encyclopedia of Health, Illness, Behavior, and Society. Wiley Online Library. 2014. Available from: https://doi.org/10.1002/9781118410868.wbehibs410.

32. Bandura A. Social cognitive theory. In P.A.M. Van Lange, A.W. Kruglanski, & E. T. Higgins (Eds.), Handbook of theories of social psychology (pp. 349 – 373). Sage Publications Ltd. 2012. Available from: https://doi.org/10.4135/9781446249215.n18.

33. World Health Organization (WHO) and Johns Hopkins Bloomberg School of Public Health. Center for Communication Programs. Information and Knowledge for Optimal Health (INFO). Decision-making tool for family planning clients and providers. Baltimore, Maryland, INFO and Geneva, WHO. (WHO Family Planning Cornerstone). 2015. [Cited 2019 12 6]. Available from: https://apps.who.int/iris/bitstream/handle/10665/43225/9241593229_eng.pdf;jsessionid=9D552FBA2D0A7F09857A01137127E5FE?sequence=2.

34. Peipert JF, Zhao Q, Allsworth JE, Petrosky E, Madden T, Eisenberg D, et al. Continuation and satisfaction of reversible contraception. Obstet. Gynecol. 2011; 117(5):1105 – 1113. Available from: https://doi.org/10.1097/AOG.0b013e31821188ad.

35. Yisa SB, Okenwa AA, Husemeyer RP. Treatment of pelvic endometriosis with etonogestrel subdermal implant (Implanon®). J Fam Plann Reprod Health Care. 2005; 31(1):67 – 70. Available from: https://doi.org/10.1783/0000000052972799.

36. Funk S, Miller MM, Mishell DR, Archer DF, Poindexter A, Schmidt J, et al. Safety and efficacy of Implanon, a single-rod implantable contraceptive containing etonogestrel. Contraception. 2005; 71(5):319 – 326. Available from: https://doi.org/10.1016/j.contraception.2004.11.007.

37. Hubacher D, Grimes DA. Non-contraceptive health benefits of intrauterine devices: a systematic review. Obstet Gynecol Surv. 2002; 57(2):120 – 128. Available from: https://doi.org/10.1097/00006254-200202000-00024.

38. Zheng SR, Zheng HM, Qian SZ, Sang GW, Kaper RF. A randomized multicenter study comparing the efficacy and bleeding pattern of a single–rod (Implanon) and a six-capsule (Norplant) hormonal contraceptive implant. Contraception. 1999; 60(1):1 – 8. Available from: https://doi.org/10.1016/S0010-7824(99)00053-0.

39. Soeprono R. Return to fertility after discontinuation of copper IUD use: a study of 55 pregnancies involving Multiload Cu-250 users among private patients in Indonesia. Adv Contracept. 1988; 4:95 – 107. Available from: https://doi.org/10.1007/BF01849510.

40. Buckshee K, Chatterjee P, Dhall GI, Hazra MN, Kodkany BS, Lalitha K, et al. Return of fertility following discontinuation of Norplant-II subdermal implants: ICMR task force on hormonal contraception. Contraception. 1995;51(4):237 – 242. Available from: https://doi.org/10.1016/0010-7824(95)00039-D.

41. Mushy SE, Shishido E, Leshabari S, Horiuchi S. Postpartum green star family planning decision aid for pregnant adolescents in Tanzania: A qualitative feasibility study. BMC Reprod Health. 2021; 18; 170. Available from: https://doi.org/10.1186/s12978-021-01216-6.

42. O’Connor AM (2010). User Manual-Decisional Conflict Scale (10 item questions format). Ottawa: Ottawa Hospital Research Institute. [Cited 2020 10 15] Available from: http://decisionaid.ohri.ca/docs/develop/User_Manuals/UM_Decisional_Conflict.pdf.

43. Lopez LM, Grey TW, Chen M, Hiller JE. Strategies for improving postpartum contraceptive use: evidence from non-randomized studies. Cochrane Database Syst Rev. 2014; 11. Available from: https://doi.org/10.1002/14651858.CD011298.pub22.

44. Mathers N, Ng CJ, Campbell MJ, Colwell B, Brown I, Bradley A. Clinical effectiveness of a patient decision aid to improve decision quality and glycemic control in people with diabetes making treatment choices: a cluster randomized controlled trial (PANDAs) in general practice. BMJ Open. 2012; 2:e001469. Available from: https://doi.org/10.1136/bmjopen-2012-001469.

45. Mullan RJ, Montori VM, Shah ND, Christianson TJH, Bryant SC, Guyatt GH, et al. The diabetes mellitus medication choice decision aid: A randomized trial. Arch Intern Med. 2009; 169(17):1560 – 1568. Available from: https://doi.org/10.1001/archinternmed.2009.293.

46. Nassar N, Roberts CL, Raynes-Greenow CH, Barratt A, Peat B. Evaluation of a decision aid for women with breech presentation at term: A randomized controlled trial. Int J Obstet and Gy. 2007; 114(3):325–33. Available from: https://doi.org/10.1111/j.1471-0528.2006.01206.x.

47. Brazell HD, O’Sullivan DM, Forrest A, Greene JF. Effect of a decision aid on decision making for the treatment of pelvic organ prolapse. Female Pelvic Med Reconstr Surg. 2015; 21(4):231–5. Available from: http://doi.org/10.1097/SPV.0000000000000149.

48. Vedasto O, Morris B, Furia FF. Shared decision-making between health care providers and patients at a tertiary hospital diabetic Clinic in Tanzania. BMC Health Serv Res. 2021; 21; 8. Available from: https://doi.org/10.1186/s12913-020-06041-4.

49. Nagle C, Gunn J, Bell R, Lewis S, Meiser B, Metcalfe S, et al. Use of a decision aid for prenatal testing of fetal abnormalities to improve women’s informed decision making: a cluster randomized controlled trial [ISRCTN22532458]. BJOG. 2008;115(3):339–47. Available from: https://doi.org/10.1111/j.1471-0528.2007.01576.x.PMID:18190370.

50. Chewning B, Mosena P, Wilson D, Erdman H, Potthoff S, Murphy A, et al. Evaluation of a computerized contraceptive decision aid for adolescent patients. Patient Educ Couns. 1999;38(3):227–39. Available from: https://doi.org/10.1016/s0738-3991(99)00014-2.

